# Estimation of the Basic Reproduction Number of SARS-CoV-2 in Bangladesh Using Exponential Growth Method

**DOI:** 10.1101/2020.09.29.20203885

**Authors:** Riaz Mahmud, H. M. Abrar Fahim Patwari

**Affiliations:** Department of Mathematics, Faculty of Mathematics and Computer Science, South Asian University, New Delhi 110021, India; Department of Applied Mathematics, Faculty of Science, Noakhali Science and Technology University, Noakhali 3814, Bangladesh

**Keywords:** Basic reproduction number, SARS-CoV-2, COVID-19, Coronavirus disease 2019

## Abstract

**Objectives:** In December 2019, a novel coronavirus (SARS-CoV-2) outbreak emerged in Wuhan, Hubei Province, China. Soon, it has spread out across the world and become an ongoing pandemic. In Bangladesh, the first case of novel coronavirus (SARS-CoV-2) was detected on March 8, 2020. Since then, not many significant studies have been conducted to understand the transmission dynamics of novel coronavirus (SARS-CoV-2) in Bangladesh. In this study, we estimated the basic reproduction number R_0_ of novel coronavirus (SARS-CoV-2) in Bangladesh.

**Methods:** The data of daily confirmed cases of novel coronavirus (SARS-CoV-2) in Bangladesh and the reported values of generation time of novel coronavirus (SARS-CoV-2) for Singapore and Tianjin, China, were collected. We calculated the basic reproduction number R_0_ by applying the exponential growth (EG) method. Epidemic data of the first 76 days and different values of generation time were used for the calculation.

**Results:** The basic reproduction number R_0_ of novel coronavirus (SARS-CoV-2) in Bangladesh is estimated to be 2.66 [95% CI: 2.58-2.75], optimized R_0_ is 2.78 [95% CI: 2.69-2.88] using generation time 5.20 with a standard deviation of 1.72 for Singapore. Using generation time 3.95 with a standard deviation of 1.51 for Tianjin, China, R_0_ is estimated to be 2.15 [95% CI: 2.09-2.20], optimized R_0_ is 2.22 [95% CI: 2.16-2.29].

**Conclusions:** The calculated basic reproduction number R_0_ of novel coronavirus (SARS-CoV-2) in Bangladesh is significantly higher than 1, which indicates its high transmissibility and contagiousness.

## Introduction

Coronavirus disease 2019 (COVID-19) is a highly infectious disease caused by severe acute respiratory syndrome coronavirus 2 (SARS-CoV-2) [1]. COVID-19 was first identified in December 2019, Wuhan, Hubei Province, China [2]. Eventually, it resulted in an ongoing pandemic [3]. Up until the 25^th^ of August 2020, more than 23.9 million cases were confirmed across 213 countries and territories. The death toll is high, with more than 820,000 deaths. Moreover, more than 16.4 million people have recovered from it [4]. However, as COVID-19 has spread out across the world, Bangladesh is not an exception. Ever since 8th March, the first case spotted, there has been a report of 299,628 confirmed cases, 4,028 deaths, and 186,756 recoveries from COVID-19 as of 25^th^ August 2020 [5]. SARS-CoV-2 infects the host’s respiratory system. It spreads between people when they come to close contact with an infected person (symptomatic or not) by inhaling small droplets produced by coughing, sneezing, or talking [6].

Most of the mathematical models on how a contagious disease spreads on a population are based on the basic reproduction number (R_0_). R_0_ defines the secondary cases generated by an infected person over his entire period of infectiousness in a completely susceptible population [7]. The significance of R_0_ lies within its threshold. If R_0_ > 1, the infected population number will grow. And if R_0_ < 1, the number will decrease [8]. In the study of epidemiology, R_0_<1 is the favorable ratio. An epidemic with high basic reproduction number R_0_ is potentially a civilization-ending threat. Several control methods can be taken to minimize the R_0_. We experienced numerous highly infectious disease outbreak in the last two decades. Globalization has enabled us with high connectivity. However, this fruit of civilization could become the doom for us instantaneously without proper control measures. We must know what we are up against to implement proper control methods. And *R*_*0*_ gives an explicit idea about the transmission of the virus.

There are many popular methods for estimating the R_0_ value. For this research purpose, we shall use the exponential growth (EG) method. Using the EG has adequacy because researchers found from the simulation that it is less prone to bias [9]. In the EG method, we need generation time (or generation interval) GT, to formulate R_0_. GT defines the time interval between symptoms recorded on an infected person and a secondary case [10].

## Materials and Methods

### Data Sources

Data used in this paper is the daily confirmed cases of infection of SARS-CoV-2 in Bangladesh (from the 1^st^ day of the outbreak to the 76^th^ day). The data were gathered from https://ourworldindata.org/coronavirus-source-data. This website provides real-time data and statistics on various topics.

### Estimation of the Basic reproduction number R_0_

The use of the basic reproduction number R_0_ is one of the most important concepts in the study of demography, ecology, and epidemiology. In epidemiology, R_0_ defines the secondary cases generated by an infected person over his entire period of infectiousness in a completely susceptible population [7]. R_0_ expresses the transmissibility of a disease. The basic reproduction number is the common buzzword during a pandemic. Because the nature of the virus is explicit with it. There are many existing methods for estimating R_0_. Among them are Exponential Growth (EG), Maximum Likelihood estimation (ML), Sequential Bayesian method (SB), the Time-Dependent method (TD), and so on. For this particular paper, we shall apply the EG method. Also, the EG method requires Generation Time (GT), also known as Generation Interval, of the virus for the estimation.

### Exponential Growth

The count of cases increases exponentially in the early stages of an epidemic. The exponential growth (EG) model is a simplified model. If something grows with a consistent rate, then it is said to be an exponential growth. The basic reproduction number R_0_ can be interpreted indirectly from the exponential curve of epidemic growth. As reported by Wallinga and Lipsitch, 2007 [11], the exponential growth of an epidemic at the early stage is linked to R_0_ as the following manner, 

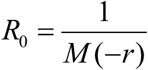

Here, M = moment generating function of GT distribution and r = exponential growth rate. The daily confirmed cases data are integers. So Poisson regression is used to fit the value of the growth rate, r.

### Generation Time

Generation Time (GT) of a virus shapes how a pandemic spread in a population. The measure of GT is critical in the estimation of R_0_. We relied upon a paper that evaluated the generation time of SARS-CoV-2. Researchers found that the GT of SARS-CoV-2 differs. For the data of Singapore, they estimated the generation time (GT) of SARS-CoV-2 to be 5.20 with a standard deviation (SD) 1.72 and the generation time (GT) 3.95 with a standard deviation 1.51 is calculated for the data of Tianjin, China [12].

### R0 Package

R software (version 4.0.2), along with the R0 package, have been used for the estimation of R_0_ and analyzing the data [9].

## Results

Using reported generation time of novel coronavirus (SARS-CoV-2) for Singapore and Tianjin, China, 5.2, and 3.95 with a standard deviation of 1.72 and 1.51 respectively, the basic reproduction number R_0_ of novel coronavirus (SARS-CoV-2) in Bangladesh are estimated. For the estimation, the data of the first 76 days of novel coronavirus (SARS-CoV-2) outbreak in Bangladesh is used. The optimal time interval is calculated to be from day 2 of the outbreak to day 44. Hence, the optimal R_0_ is calculated in the optimal time interval.

The value of R_0_ is estimated to be 2.66 [95% CI: 2.58-2.75] with the optimal value of 2.78 [95% CI: 2.69-2.88], using the reported generation time of SARS-CoV-2 for Singapore. Furthermore, using the reported generation time of SARS-CoV-2 for Tianjin, China, the estimated R_0_ is 2.15 [95% CI: 2.09-2.20] with the optimal value of 2.22 [95% CI: 2.16-2.29] (Table 1).

**Table 1.** Estimation of basic reproduction number R_0_ of SARS-CoV-2 in Bangladesh by exponential growth method using different reported values of generation time of SARS-CoV-2.

| Generation Time | Source | Remark | Estimated $R_0$<br>[95% CI] | Optimal $R_0$<br>[95% CI] |
| --- | --- | --- | --- | --- |
| GT 5.20, SD 1.72 | [12] | GT was reported for Singapore. | 2.66<br>[2.58-2.75] | 2.78<br>[2.69-2.88] |
| GT 3.95, SD 1.51 | [12] | GT was reported for Tianjin, China. | 2.15<br>[2.09-2.20] | 2.22<br>[2.16-2.29] |

### Sensitivity Analysis

To analyze the sensitivity, we estimated R_0_ of SARS-CoV-2 for the same data using previously reported generation time of MERS and SARS coronavirus 7.6 and 8.4 with a standard deviation of 3.4 and 3.8, respectively [13, 14]. The estimated R_0_ is 3.65 [95% CI: 3.51-3.80] and 4.07 [95% CI: 3.90-4.25] with optimal value 3.86 [95% CI: 3.70-4.04] and 4.32 [95% CI: 4.13-4.53] corresponding to the generation time of MERS and SARS coronavirus respectively (Table 2).

**Table 2.**
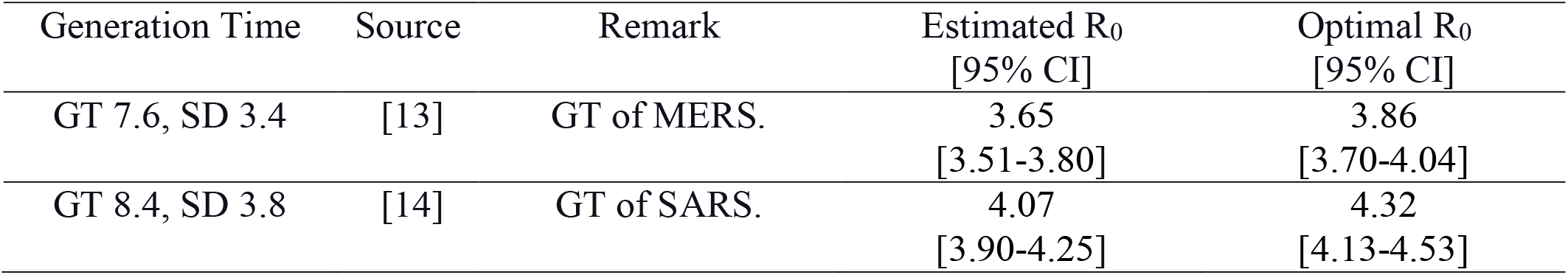
Estimation of basic reproduction number R_0_ of SARS-CoV-2 in Bangladesh by exponential growth method using generation time of MERS and SARS coronavirus.

We found that the value of R_0_ varies with the change of generation time. Using the EG method, for generation time 3.95, the value of R_0_ is estimated to be 2.15, and this value is almost doubled and estimated to be 4.07 for generation time 8.4 (Figure 1).

**Figure 1.**
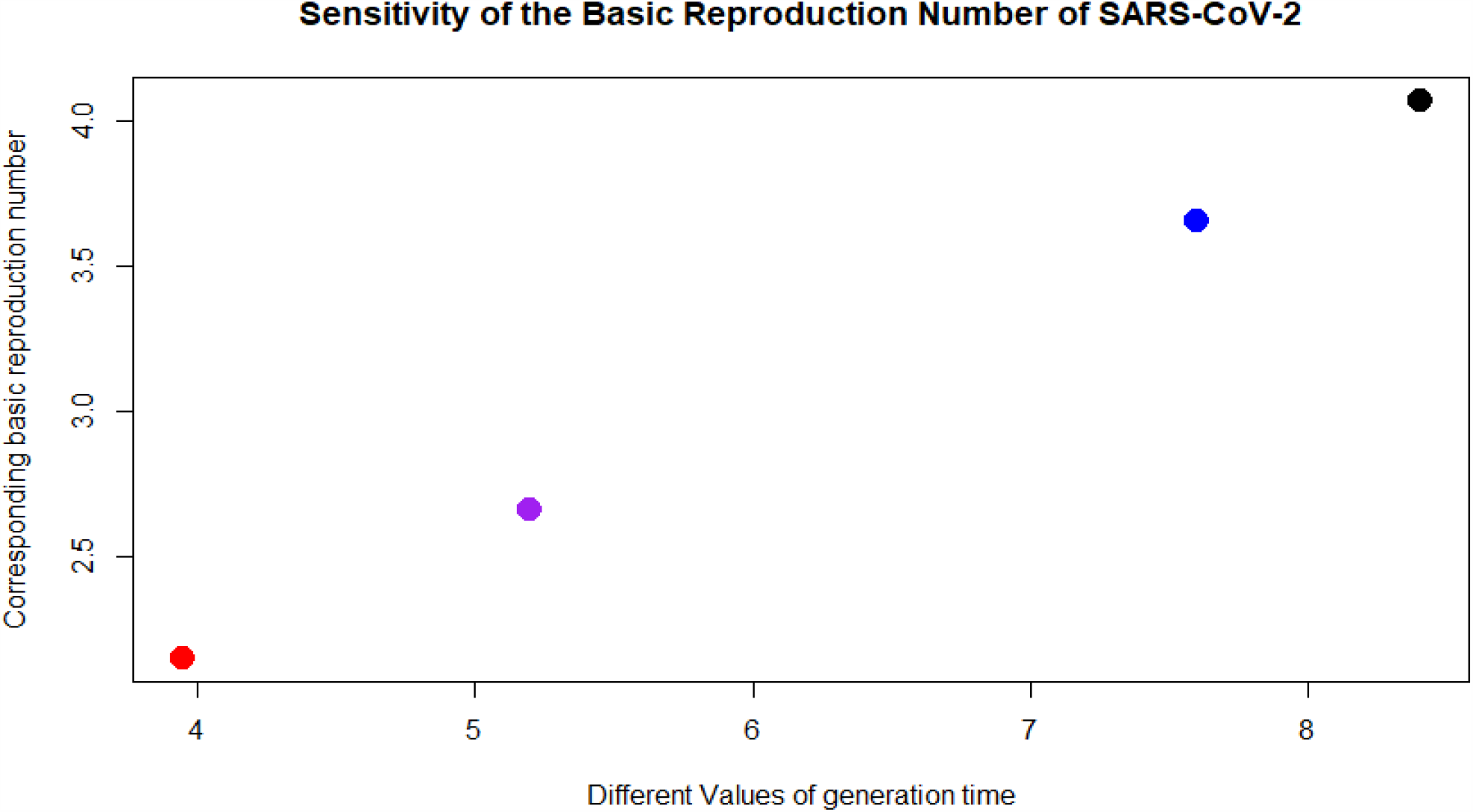
Different values of R_0_ corresponding to different values of generation time is illustrated. The values of R_0_ corresponding to generation time 3.95, 5.20, 7.60, and 8.40 represented by red, purple, blue, and black dots, respectively.

The analysis shows that R_0_ is sensitive to generation time. Hence, generation time needs to be estimated accurately to estimate R_0_ more precisely.

## Discussion

The estimated value of the basic reproduction number R_0_ of SARS-CoV-2 in Bangladesh using the exponential growth method is ranging from 2.22 to 2.78. This result is quite similar to the previously reported R_0_ using the same method in China 2.24 and 2.90 [15, 16]. This estimated value of R_0_ is significantly greater than 1, which is evidence of the high transmissibility and contagiousness of SARS-CoV-2 in Bangladesh. In this study, we found that R_0_ shows some sensitivity to generation time. Thus, a more precise estimation of generation time is needed to estimate R_0_ with more accuracy.

The estimation of R_0_ can be used to estimate other transmission dynamical parameters of SARS-CoV-2 in Bangladesh. It also can help to determine an optimum strategy for vaccination. From the epidemiological study, we have,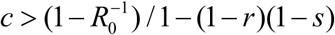 [17]. Where c is the minimum coverage of elimination, r is the fraction of individuals who are completely immunized, and s is the susceptible individuals.

This study has some strong points. First, in this study the quantification of the basic reproduction number R_0_ of SARS-CoV-2 in Bangladesh has been done using the reported generation time of SARS-CoV-2. This increases the possibility of better accuracy of the study. Second, the sensitivity analysis was done for different generation time values, which gives a hint for further betterment of the estimation of R_0_. Last, this study gives a hint for the optimal vaccination strategy. This can be helpful if the vaccine arrives in the near future.

This study has several limitations too. First is the quality of data. We collected data from a public resource. The quality of data is not ensured. There is a possibility that some confirmed cases remained unreported. This can affect our estimation. Second, the reported values of generation time vary from 3.95 to 5.20. This is not quantified precisely.

In summary, using the exponential growth method, the basic reproduction number R_0_ of SARS-CoV-2 is estimated. R_0_ has shown some sensitivity to generation time. This study can be helpful for further understanding of the transmission dynamics of SARS-CoV-2 in Bangladesh. The estimated R_0_ is significantly larger than 1, which is evidence of its high transmissibility and contagiousness. Either vaccination should be done, or more preventive control measures should be taken to reduce the epidemic size.

## Data Availability

In this study, the data were collected from publicly available resources.

https://ourworldindata.org/coronavirus-source-data

https://www.worldometers.info/coronavirus/

## References

1. World Health Organization. Naming the coronavirus disease (COVID-19) and the virus that causes it. [Online URL: https://www.who.int/emergencies/diseases/novel-coronavirus-2019/technical-guidance/naming-the-coronavirus-disease-(covid-2019)-and-the-virus-that-causes-it] accessed on August 25, 2020.

2. World Health Organization. Novel Coronavirus – China. [Online URL: https://www.who.int/csr/don/12-january-2020-novel-coronavirus-china/en/] accessed on August 25, 2020.

3. World Health Organization. WHO Director-General’s opening remarks at the media briefing on COVID-19 - 11 March 2020. [Online URL: https://www.who.int/dg/speeches/detail/who-director-general-s-opening-remarks-at-the-media-briefing-on-covid-19---11-march-2020] accessed on August 25, 2020.

4. Worldometer.info. COVID-19 Coronavirus Pandemic. [Online URL: https://www.worldometers.info/coronavirus/] accessed on August 25, 2020.

5. Worldometer.info. Bangladesh Coronavirus cases. [Online URL: https://www.worldometers.info/coronavirus/country/bangladesh/] accessed on August 25, 2020.

6. World Health Organization. Q&A on coronaviruses (COVID-19). [Online URL: https://www.who.int/emergencies/diseases/novel-coronavirus-2019/question-and-answers-hub/q-a-detail/q-a-coronaviruses] accessed on August 25, 2020.

7. Fraser, C., Donnelly, C. A., Cauchemez, S., Hanage, W. P., Van Kerkhove, M. D., Hollingsworth, T. D., et al. (2009). Pandemic potential of a strain of influenza A (H1N1): early findings. Science (New York, N.Y.), 324, 1557–1561.

8. Heesterbeek, J. A. (2002). A brief history of R0 and a recipe for its calculation. Acta Biotheor, 50, 189–204.

9. Obadia, T., Haneef, R., Boëlle, P. Y. (2012). The R0 package: a toolbox to estimate reproduction numbers for epidemic outbreaks. BMC Med Inform Decis Mak, 12, 147.

10. Park, S. W., Champredon, D., Weitz, J. S., Dushoff, J. (2019). A practical generation-interval-based approach to inferring the strength of epidemics from their speed. Epidemics, 27, 12–18.

11. Wallinga, J., Lipsitch, M. (2007). How generation intervals shape the relationship between growth rates and reproductive numbers. Proceedings. Biological sciences / The Royal Society, 274, 599–604.

12. Ganyani, T., Kremer, C., Chen, D., Torneri, A., Faes, C., Wallinga, J., et al. (2020). Estimating the generation interval for coronavirus disease (COVID-19) based on symptom onset data, March 2020. Euro surveillance : bulletin Europeen sur les maladies transmissibles = European communicable disease bulletin, 25, 2000257.

13. Assiri, A., McGeer, A., Perl, T. M., Price, C. S., Al Rabeeah, A. A., Cummings, D. A., et al. (2013). Hospital outbreak of Middle East respiratory syndrome coronavirus. N Engl J Med, 369, 407–416.

14. Lipsitch, M., Cohen, T., Cooper, B., Robins, J. M., Ma, S., James, L., et al. (2003). Transmission dynamics and control of severe acute respiratory syndrome. Science (New York, N.Y.), 300, 1966–1970.

15. Zhao, S., Lin, Q., Ran, J., Musa, S. S., Yang, G., Wang, W., et al. (2020). Preliminary estimation of the basic reproduction number of novel coronavirus (2019-nCoV) in China, from 2019 to 2020: A data-driven analysis in the early phase of the outbreak. Int J Infect Dis, 92, 214– 217.

16. Liu, T., Hu, J., Kang? M., Lin, L., Zhong, H., Xiao, J., (2020). et al. Transmission dynamics of 2019 novel coronavirus (2019-nCoV). bioRxiv, [Online URL: http://dx.doi.org/10.2139/ssrn.3526307] accessed on August 25, 2020.

17. Dietz, K. (1993). The estimation of the basic reproduction number for infectious diseases. Statistical methods in medical research, 2, 23–41.

